# Insular Lesions as a Neural Substrate of Attentional Dysfunction in Post-Stroke Depression

**DOI:** 10.64898/2025.12.17.25342446

**Authors:** Yoonhye Na, Dorothee P. Auer, JeYoung Jung, Sung-Bom Pyun

**Author notes:** **Corresponding authors** Sung-Bom Pyun, Department of Physical Medicine and Rehabilitation, Korea University Anam Hospital, 73 Goryeodae-ro, Seongbuk-gu, Seoul 02841, Republic of Korea, JeYoung Jung, School of Psychology, University of Nottingham, Nottingham, UK.

## Abstract

Post-stroke depression (PSD) represents one of the most prevalent psychiatric complications after stroke, affecting up to one-third of survivors, and it can substantially influence both the extent of functional impairments and the recovery trajectory. A reciprocal relationship between depressive symptoms and cognitive dysfunction in this population has been commonly demonstrated, underscoring the need to clarify their intercorrelation and shared neuroanatomical underpinnings. To address this a combined cognitive dysfunction linked to PSD and imaging study, a total of 125 unilateral stroke patients were classified into two groups based on the presence of depressive symptoms using a BDI cutoff score of 14. Using principal component analysis (PCA) on 16 cognitive test variables from 64 patients with PSD and 61 without PSD, four principal components of cognitive function were extracted. Among these, the attention factor showed a significant group difference, with the PSD group exhibiting greater attentional impairments than the non-PSD group. Voxel-based lesion-symptom mapping (VLSM) further revealed that attentional performance was significantly associated with lesions in the left insula across groups. Importantly, the lesion clusters were more extensive in patients with PSD, suggesting a more pronounced role of insular damage in depressive symptomatology following stroke. Taken together, these findings highlight the pivotal contribution of the insula to both attentional processing and the pathophysiology of PSD, suggesting that the insula mediates attentional networks closely linked to depressive symptoms in stroke survivors.

## Introduction

Post-stroke depression (PSD) is one of the most common psychiatric complications following a stroke, affecting up to one-third of stroke survivors ^1^. It is closely linked to the extent of functional impairments and the recovery trajectory ^2-5^. Neurological and cognitive disorders such as aphasia, dysarthria, or dysphagia significantly contribute to the development of depressive symptoms, likely due to increased social isolation and frustration from communication barriers ^6^. PSD often presents as persistent sadness and loss of motivation, which can hinder rehabilitation of patients ^7^.

The bidirectional relationship between depressive symptoms and cognitive dysfunction after stroke has been repeatedly reported, with each condition exacerbating the other ^6,8-10^. Stroke patients with more severe depression are up to three times more likely to exhibit cognitive impairments compared to those with milder symptoms ^11^. Chronic depression has been identified as a key predictor of post-stroke cognitive deficits ^12^. Among the affected domains, executive function is frequently impaired, alongside deficits in memory, attention, language, non-verbal problem solving, and processing speed, etc. ^6,11,13^. These cognitive impairments may reduce patients’ ability to cope with stroke-related challenges, thereby contributing to or worsening depression.

In major depressive disorder (MDD), cognitive and emotional dysfunctions are closely linked ^14-16^ and are thought to be partly driven in the insula abnormalities ^17-19^. The insular cortex act as a hub connecting subcortical regions with various cortical areas, playing a key role in integrating sensory information and supporting higher cognitive functions ^20^. Structural and functional alterations in the insula such as decreased grey matter volume (GMV) and disrupted functional connectivity (FC) have been implicated in the pathophysiology of numerous psychiatric and neurological disorders ^21,22^. Notably, the left dorsal granular insular has been found to show reduced volume in people with MDD compared to healthy controls ^23^, and the fronto-striatal salience network was expanded in depressed populations ^24^. Furthermore, FC to the dorso-medial insular cortex (dmIC) has been associated with depression severity in MDD ^25^.

Given the high heterogeneity in lesion location, size, and symptoms presentation in stroke, identifying neural correlates of PSD remains challenging. However, recent studies have begun to reveal relevant patterns: for example, increased FC between anterior insula and superior frontal regions has been reported in PSD (Park et al., 2021), decreased insula volume has also been found in PSD patients ^26^, and even a prominent role of right insula in PSD ^27^. Despite these finding, there is still no consensus regarding the neuroanatomical basis of PSD, particularly in relation to the cognitive impairments that contributing to depressive symptom after stroke.

Understanding of the neuroanatomical correlates of PSD, especially those linked with cognitive dysfunction, holds significant clinical relevance. It may inform more targeted rehabilitation strategies and improve quality of life for both stroke survivors and their caregivers. Here, this study aims to identify the neural correlates of PSD and examine their association with cognitive deficits. The present study extracts principal components from comprehensive set of language and cognitive test variables to identify core cognitive domains that influence PSD, and elucidates the associated neural correlates, with particular focus on the insular cortex. We hypothesize that a cognitive dysfunction in PSD compared to non-depressive group is associated with the neural correlates specifically in the subregions of insula.

## Materials and methods

### Subjects

We retrospectively reviewed data from the Stroke Outcome Prediction database (STOP DB) by the department of physical medicine and rehabilitation at Korea University Anam Hospital from August 2012 to June 2023. The inclusion criteria were as follows: (i) 1^st^-ever stroke (ischemic or hemorrhagic), (ii) right-handedness, (iii) aged between 20 and 85 years, and (iv) who complete magnetic resonance (MR) imaging and behavioral assessments, including the Beck’s depression inventory (BDI) ^28^ and the computerized neurocognitive function test (CNT) battery. We also excluded patients who (i) have infra-tentorial lesions, (ii) incomplete behavioral assessments, or (iii) have medical history of psychiatric disease or neurodegenerative disease. A total 125 patients with stroke were selected from the STOP DB.

This study was approved by the Institutional Review Board (IRB) committee at Korean University Anam Hospital (IRB no. 2025AN0050).

### Behavioral assessments

All the participants completed the BDI and CNT. The BDI is a widely used self-report measure for assessing depressive symptoms, consisting of 21 items scored on a scale from 0 to 3, with total scores ranging from 0 to 63 ^28^. Higher scores indicate greater severity of depressive symptoms. Based on BDI scores, participants were categorized into two groups: Post-stroke depression (PSD) group: BDI score > 14 points and Post-stroke (PS) group: BDI score ≤ 14 points. Depressive symptoms can be clinically categorized into four levels using BDI, and previous studies have applied various cutoffs for research ^12,29,30^. In the present study, a cutoff of 14, corresponding to mild depressive symptoms, was adopted to emphasize the presence of depression ^31^. This classification yielded 64 patients in the PSD group and 61 in the PS group.

The CNT is a computerized neurocognitive test battery designed to examine a broad range of cognitive functions ^32^. It includes digit span test (DST), visual span test (VST), auditory and visual continuous performance test (aCPT and vCPT, respectively) for attention, verbal learning test (VLT) for verbal memory, Stroop test (STR) for executive function, trail making test (TMT) for mental flexibility and coloured progressive metrices (CPM) for non-verbal reasoning.

Test instructions were uniformly delivered by the software across all administrations, and participants completed several practice trials prior to the commencement of the main test. The DST and VST were employed to evaluate short term memory function by requiring participants to recall sequences of numbers or visual stimuli either in the order presented (forward) or in reverse (backward). The CPT was used to assess automatic and selective attention through simple detection rules. In the aCPT, participants pressed a button as quickly as possible upon hearing the word “three”, whereas in the vCPT, they responded to the appearance of the number “3” among sequentially presented numbers. The VLT evaluated verbal memory, and participants listened to a list of 15 recorded words and attempted to recall as many as possible across five learning trials. Among derived scores of the VLT, we used the score of the first list as an immediate memory and total score for the verbal memory span. The STR consisted of five conditions. STR1 involved reading words printed in black, representing an automatic reading process. STR2 required naming the colors of colored squares. In STR3 and STR4, participants read colored words with congruent (identical) or incongruent (non-identical) word–color pairings, the latter demanding greater inhibition of distraction. STR5 required naming the color of the word when the written word and the ink color were incongruent, further increasing inhibitory demands. The TMT included two subtests. TMT_A required participants to sequentially click numbers from 1 to 25, while TMT_B involved alternating responses between numbers and Hangeul characters in an ordered sequence (e.g., pressing the number “1,” then the Hangeul character “가 (/ga/),” followed by the number “2,” then “나 (/na/),” and so on). The CPM required participants to select the correct stimulus based on perceptual features and logical judgment. All the tests have the standardized norms by age, sex, and education ^32-34^. We used the score or percentile of each test for further statistical analysis as a variable reflecting the participants’ performance.

### Statistical analyses

To reduce dimensionality and identify underlying components in the behavioral data, we employed Principal Component Analysis (PCA), a data-driven multivariate statistical technique ^35^. This approach enables the summarization of a large set of correlated variables into a smaller number of uncorrelated principal components, capturing the core structure of the dataset. In this study, PCA was conducted on the cognitive test scores to extract latent cognitive domains, which were subsequently used as composite cognitive function variables in further analyses. We used raw scores or percentiles of following tests: (i) digit span test (forward/backward), (ii) visual span test (forward/backward), (iii) auditory continuous performance test (correct response), (iv) visual continuous performance test (correct response), (v) verbal learning test (immediate recall and total score), (vi) Stroop test (percentile), (vii) trail making test (number only/number and letter), and (viii) colored progressive matrices (total score). PCA was performed using varimax rotation to improve interpretability of the factor structure. Variables with factor loadings ≥ 0.50 on the same component were grouped together and interpreted as representing a common cognitive domain. These extracted components were then used as composite cognitive scores in subsequent analyses examining the relationship between cognitive performance and post-stroke depression.

Independent sample t-tests were used to examine patients’ demographic characteristics, and chi-square tests were applied to categorical variables such as sex and lesion side. All statistical analyses were conducted using IBM SPSS Statistics (Version 22.0).

### MRI acquisition

All patients underwent MRI scanning after the onset of stroke. We collected the high-resolution structural T1-weighted MRI and diffusion tensor image (DTI). All MR images were acquired on a 3.0T Siemens Prisma scanner (Siemens, Erlangen, Germany) at Korea University Anam Hospital.

A T1-weighted image was acquired with the following parameters: repetition time (TR) = 2020 ms, echo time (TE) = 2.91 ms, flip angle = 9º, filed of view (FOV) = 217 × 166 mm^2^, voxel size = 0.34 × 0.8 × 0.34 mm^3^. A DTI was acquired with 64 diffusion gradient directions and the following acquisition parameters: repetition time (TR) = 6500 ms, echo time (TE) = 55 ms, flip angle = 90º, field of view (FOV) = 224 x 224 mm^2^, voxel size = 1.0 x 1.0 x 2.0 mm^3^, b = 1000 sec/mm^2^.

### Voxel-based lesion symptom mapping (VLSM)

Lesion segmentation was conducted manually using ITK-SNAP ver. 3.8.0 ^36^, and the procedure was conducted in a blinded manner with respect to the participants’ PSD status. Lesions were identified on the B0 image, the first volume of diffusion weighted image (DWI). Binary lesion masks were then coregistered to each individual T1-weighted anatomical image using Statistical Parametric Mapping (SPM) 12 (https://www.fil.ion.ucl.ac.uk/spm/software/spm12), and normalized to the MNI 152 template. Statistical models were built using Statistical Non-Parametric Mapping (SnPM) 13 toolbox implemented in MATLAB (version R2023a, The MathWorks, Inc., Natick, Massachusetts, United States). For the voxel-based lesion-symptom mapping (VLSM) analysis, four cases of PS and five cases of PSD were excluded due to bilateral or multi focal lesions. Statistical model was built using a two-sample t-test to compare two groups. This statistical model created analysis design and permutation matrix appropriate for two group comparison where there was one scan per subject. Statistical significance was determined using permutation testing with 5,000 iterations. To control potential confounds, age, sex, years of education, and lesion volume were included as covariates in the VLSM model. Results that survived family-wise error (FWE) correction at p < 0.05 were reported.

In order to investigate the specific contribution of insula subregions to PSD, we conducted a regions of interest (ROI) analysis using the Julich Brain Atlas framework ^37^. This high-resolution cytoarchitectonic atlas provides detailed parcellation based on microstructural and connectivity data. We defined sixteen subregions of insula from each hemisphere, comprising three agranular insular cortices (IA), ten dysgranular insular cortices (ID), and three granular cortices (IG) as ROIs.

### Lesion overlay map

Lesion overlay maps for the PS and PSD groups are shown in Figure 1. In both groups, the highest overlap of lesions was observed in regions centered around the basal ganglia.

**Figure 1.**
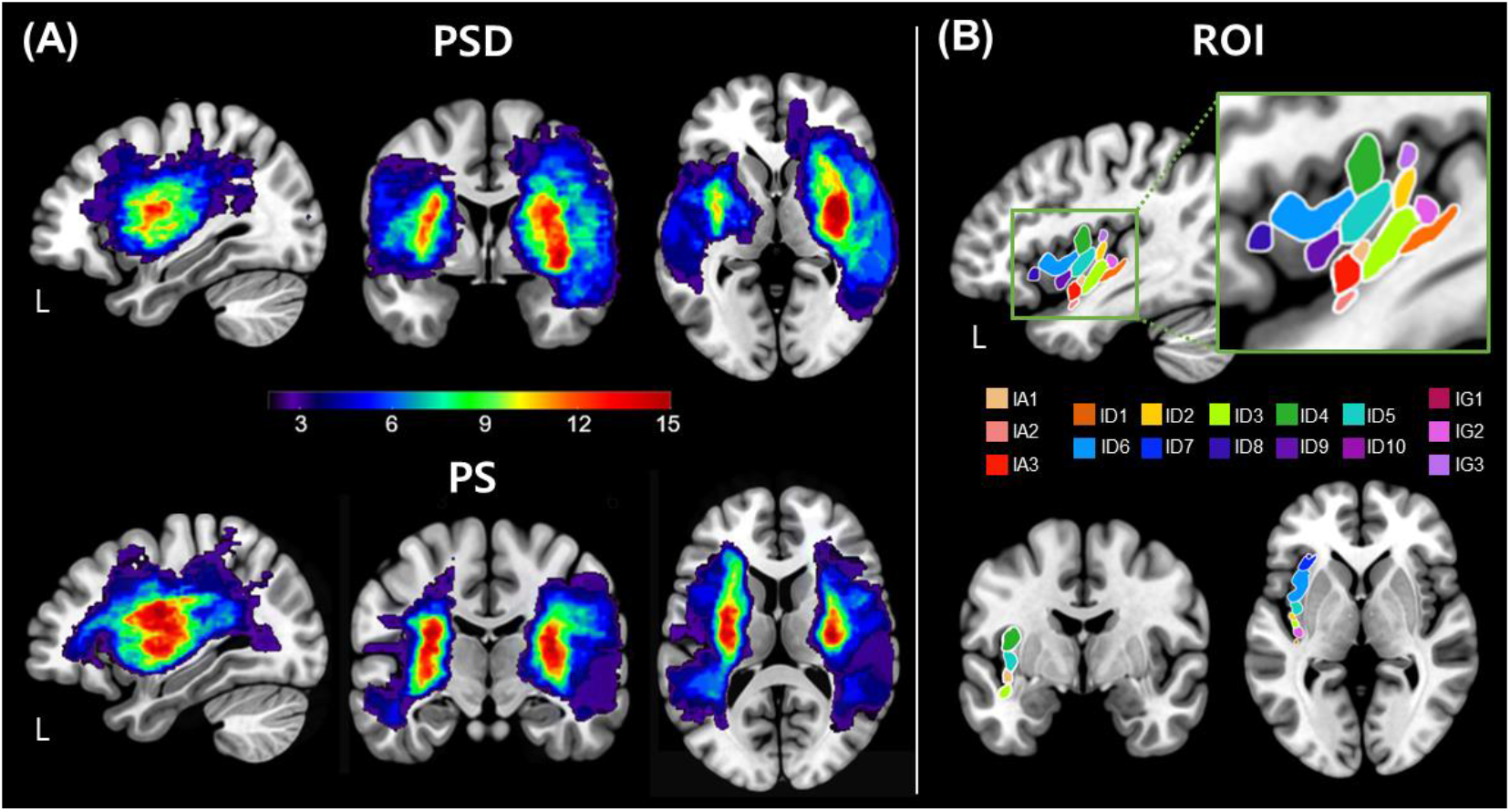
Lesion overlay maps and ROIs. (A) Lesion overlap maps for the post-stroke (PS) and post-stroke depression (PSD) groups. Warmer colors represent regions with greater lesion overlap across patients. The color bar indicates the number of patients with overlapping lesions at each voxel. (B) ROIs in the insula subregions

## Results

### Demographic results

The demographic and clinical characteristics of the participant are summarised in Table1. The mean age of PSD group was 59.97 years (SD = 14.13), and that of PS group was 57.57 years (SD = 13.97). There were no significant differences between the two groups in terms of age, sex, years of education, and lesion laterality. Cognitive function, as measured by the Korean Mini-Mental State Examination (K-MMSE), did not differ significantly between the groups. However, BDI scores were significantly higher in the PSD group compared to the PS group, indicating greater depressive symptoms (t(128) = −14.805, p <.001). The lesion volume was not statistically different (t(128)=-1.209, p=.229). The timing of MRI scanning and the cognitive and depression assessments conducted, on average, one month after stroke onset, and no statistically significant differences were observed between the groups.

**Table 1.** The demographic and clinical characteristics of the PSD and PS groups.

|  | **PSD**<br>(N=64) | **PS**<br>(N=61) | **T/ $\chi^2$** | **P** |
| --- | --- | --- | --- | --- |
| **Age (yr)** | 59.91<br>(14.13) | 57.57<br>(13.97) | -.928 | .355 |
| **Education (yr)** | 11.42<br>(4.38) | 12.69<br>(3.72) | 1.740 | .084 |
| **MMSE (30)** | 22.23<br>(6.33) | 21.49<br>(8.93) | -.538 | .591 |
| **BDI (63)** | 24.34<br>(8.73) | 6.93<br>(3.64) | -14.424 | <.001<br>* |
| **Sex (M/F)** | 37 / 27 | 36 / 25 | 0.019 | .981 |
| **Lesion side (L/R)** | 31 / 33 | 40 / 21 | 3.738 | .053 |
| **Lesion volume (mm<sup>3</sup>)** | 43909.33<br>(50044.33) | 33195.91<br>(43331.88) | -1.209 | .229 |
| **MRI post onset (day)** | 24.23<br>(35.01) | 44.58<br>(115.00) | 1.276 | .204 |
| **Test post onset (day)** | 17.78<br>(30.23) | 13.76<br>(22.21) | -.800 | .425 |

### PCA results

Principal Component Analysis (PCA) was conducted on 16 variables derived from the cognitive function tests. The analysis yielded four independent components, which accounted for 71.83% of the total variance (factor 1 = 20.42%, factor 2 = 20.03%, factor 3 = 16.30%, and factor 4 = 15.09%) (Table 2 and Figure 2). The factor loadings for each variable are shown in Table 2. Variables with loadings ≥ 0.50 were grouped under the same factor. Based on the nature of the loaded variables, the factors were interpreted and labeled as follows: Factor 1:verbal memory, Factor 2:cognitive control, Factor 3:attention, and Factor 4:mental flexibility

**Table 2.** Principal Component Analysis (PCA) factor loadings for cognitive test variables.

|  | Principal components |  |  |  |
| --- | --- | --- | --- | --- |
|  | Verbal memory | Cognitive Control | Attention | Mental flexibility |
| **VLT list 1 score** | **.909** | .055 | .181 | .001 |
| **VLT total score** | **.903** | -.010 | .219 | .084 |
| **DST_B** | **.823** | .138 | .184 | .214 |
| **DST_F** | **.755** | .058 | .185 | .293 |
| **STR 3** | .099 | **.881** | .000 | .107 |
| **STR 2** | .050 | **.864** | .064 | .051 |
| **STR 1** | .216 | **.757** | .169 | .230 |
| **STR 5** | -.061 | **.699** | .106 | -.058 |
| **STR 4** | -.004 | **.667** | .017 | .340 |
| **VCPT** | .169 | .228 | **.806** | -.029 |
| **VST_B** | .062 | .096 | **.722** | .485 |
| **ACPT** | .296 | .012 | **.710** | .024 |
| **VST_F** | .218 | -.009 | **.603** | **.556** |
| **CPM** | .298 | .062 | **.583** | .403 |
| **TMT_B** | .182 | .216 | .139 | **.831** |
| **TMT_A** | .162 | .195 | .134 | **.831** |

\* Factor loadings over 0.5 were presented in bold
\*\* abbreviations. VLT; verbal learning test, SC; score, T\_SC; total score, DST\_B; digit span test (backward), DST\_F; digit span test (forward), STR; Stroop test, VCPT; visual continuous performance test, CR; correct response, VST\_B; visual span test (backward), ACPT; auditory continuous performance test, VST\_F; visual span test (forward), CPM; coloured progressive matrices, TMT; trail making test

**Figure 2.**
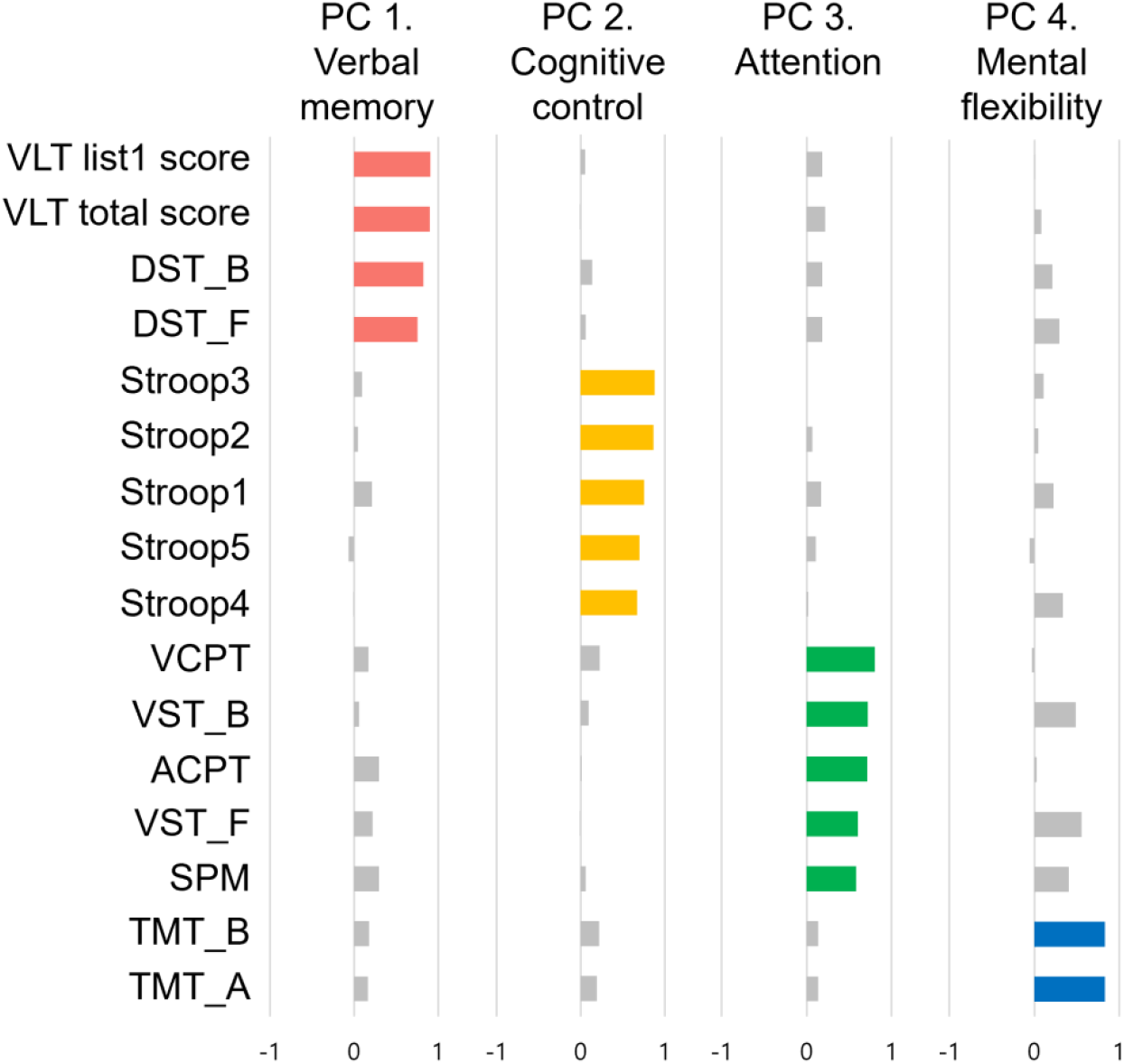
Principal components and the factor loadings

A group comparison revealed a significant difference between the PSD and PS groups on Factor 3 (Attention), with the PSD group showing lower performance [t(123) = 2.153, p =.034]. (Figure 3 - b).

**Figure 3.**
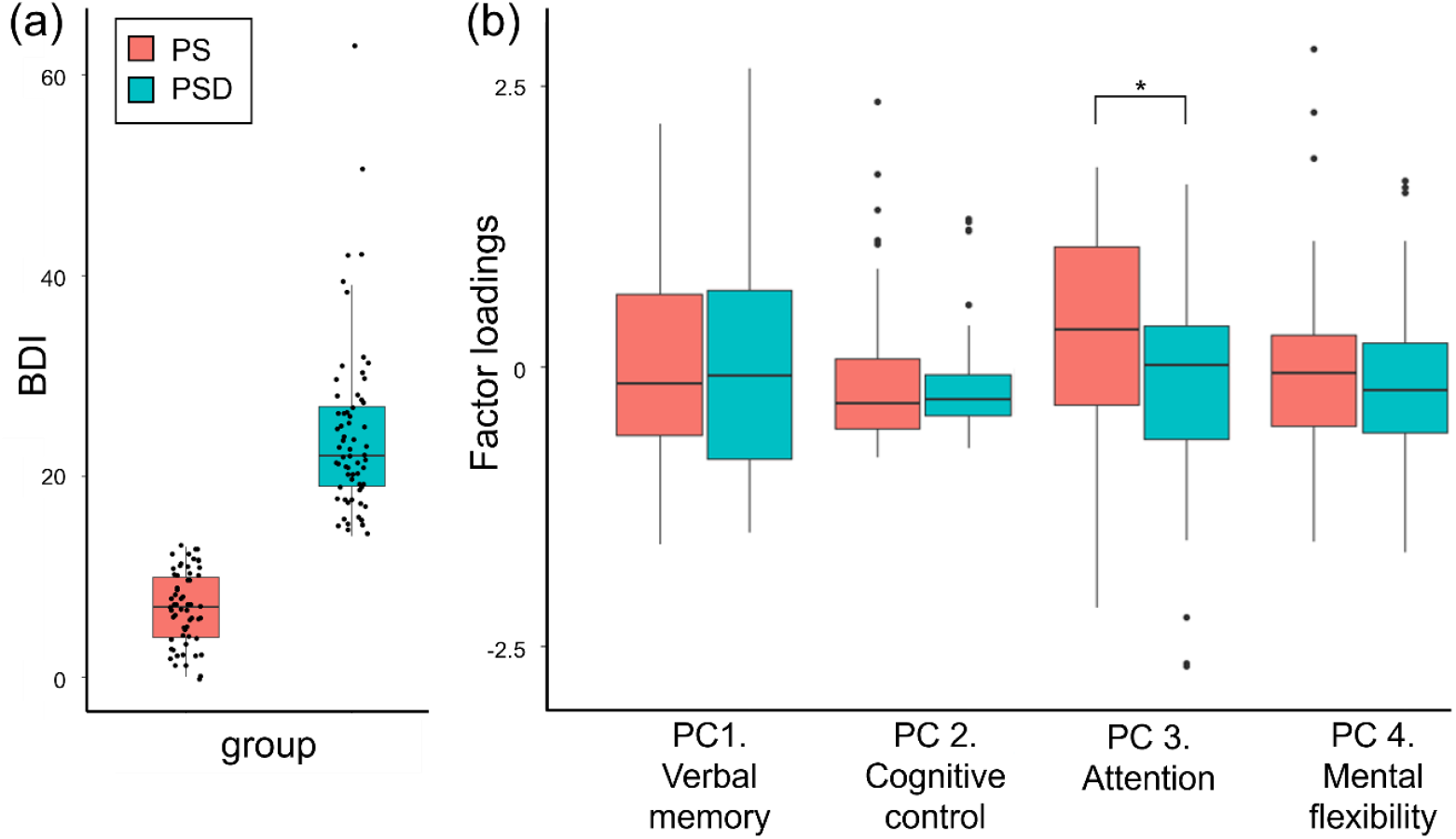
BDI scores (a) and Factor loadings (b) by group

### VLSM results

VLSM result showed that there was no significant direct relationship between lesion and BDI score in both groups. The relationships between lesion location and factor scores showed a similar pattern in both the PSD and PS groups (Figure. 4). In both groups, a significant relationship was observed between factor 3 (attention) and lesions in the left insula. Additionally, the PS group showed a significant relationship between factor 3 and lesion in the left middle temporal gyrus (MTG). The lesion cluster in the left insula was larger in the PSD group compared to the PS group, with a small area of overlap observed within the left insular cortex across both groups. No significant between-group differences were identified in the whole-brain analysis.

**Figure 4.**
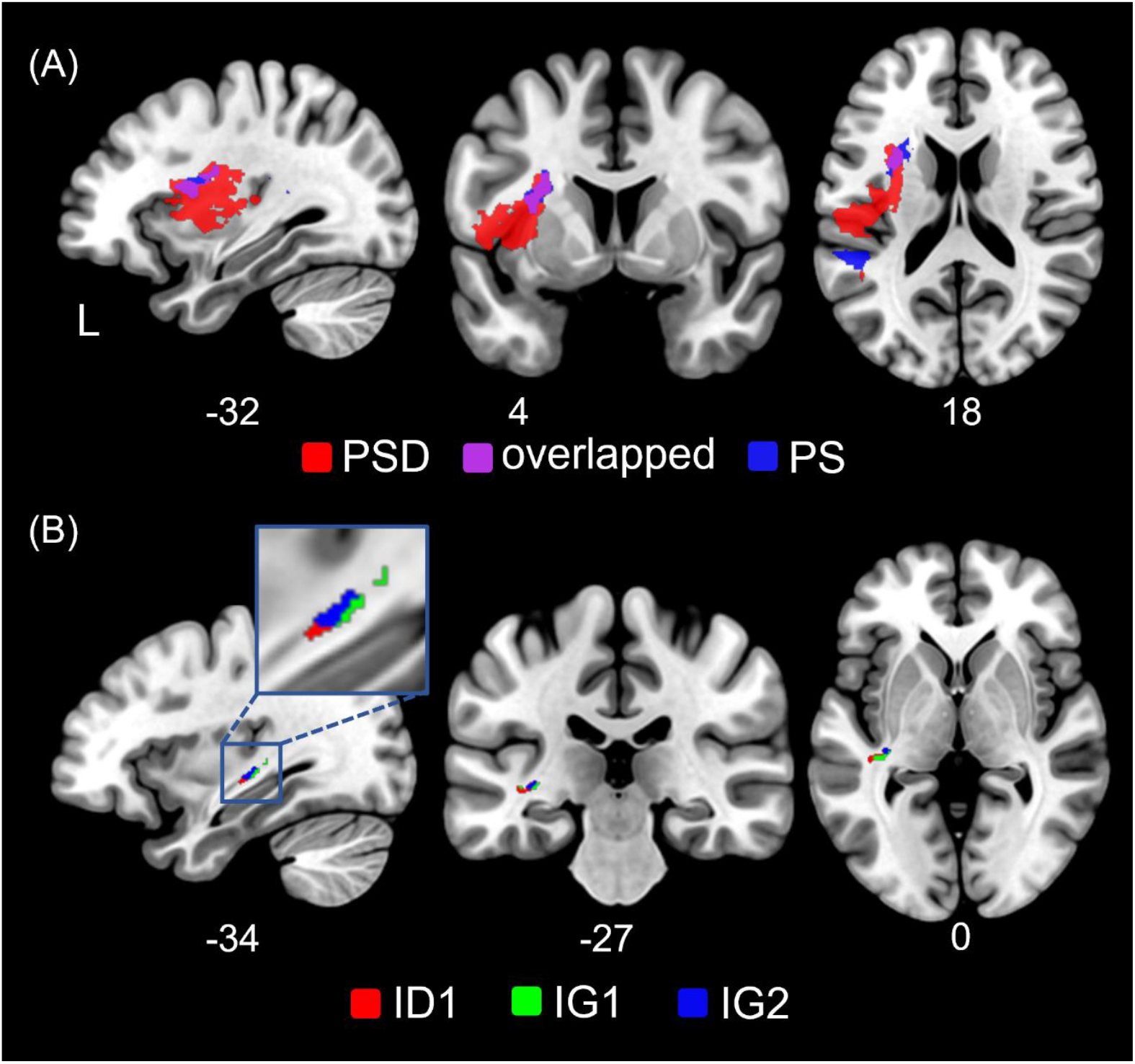
VLSM results. (A) Neuroanatomical correlates of factor 3 in PS (red) and PSD (blue) and (B) ROI analysis results. PSD: post-stroke depression, PS: post-stroke, ID: dysgranular insular cortex, IG; granular insular cortex

The ROI analysis revealed group differences in the posterior left insula (Figure 4 and Table 3). Specifically, the PSD group showed more lesion overlaps compared to the PS group in the dysgranular insular cortex 1 (ID1) (p = 0.048). And there was marginal difference in the granular insular cortices 1 (IG1) (p = 0.053) and 2 (IG2) (p = 0.064).

**Table 3.**
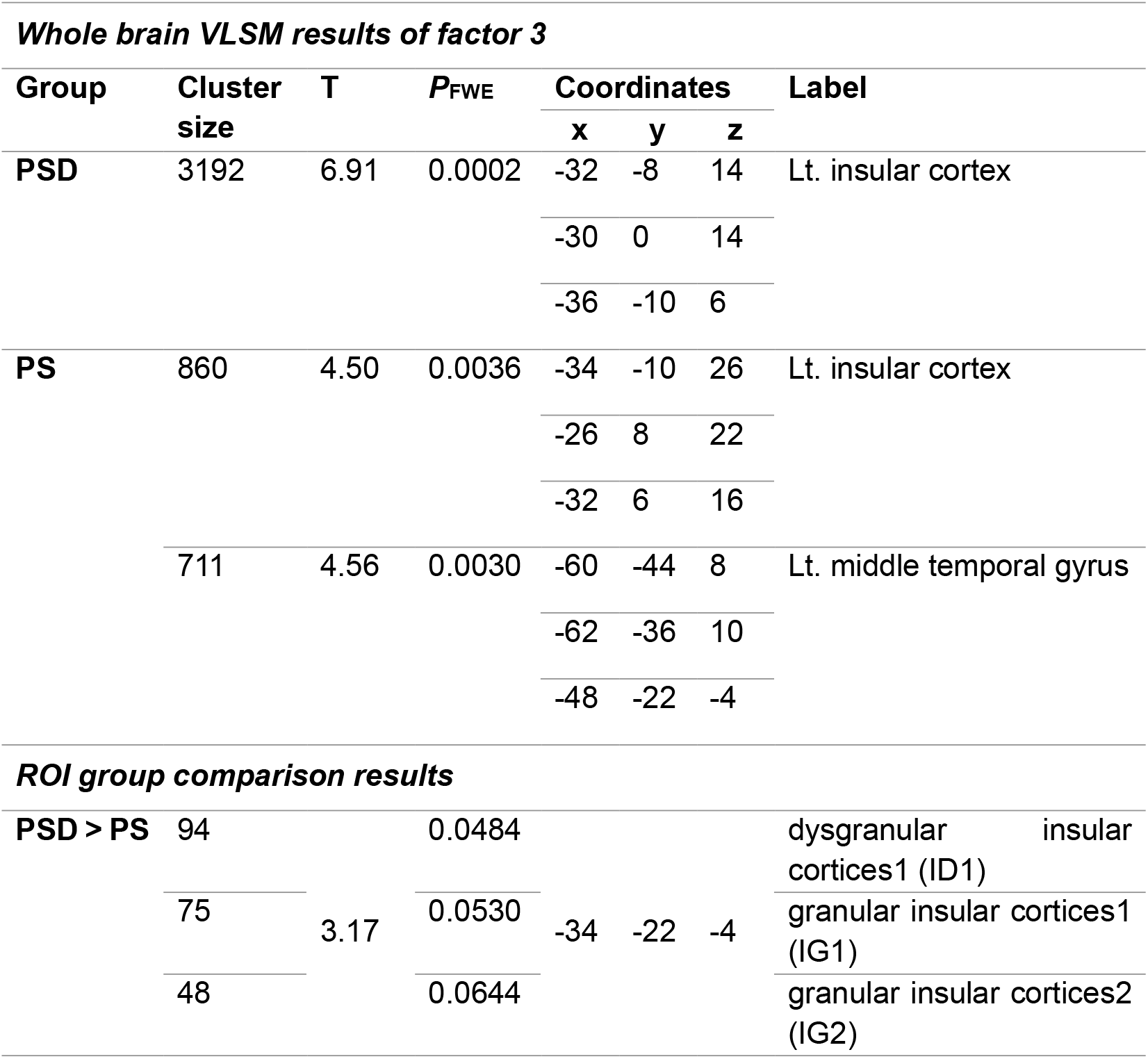
VLSM results and ROI analysis results for attention factor.

## Discussion

This study examined the cognitive correlates of PSD and their underlying neuroanatomical substrates. Using PCA of 16 cognitive test variables, we identified four principal components of cognitive function. Among these, the attention factor showed a significant group difference, with PSD group exhibiting greater attentional deficits than the PS group. VLSM further revealed that the attention factor was significantly associated with lesions in the left insula in both groups. Notably, the lesion cluster was more extensive in the PSD group than in the PS group, suggesting a more pronounced role of insular damage in patients experiencing depressive symptoms after stroke. Together, these findings highlight the critical contribution of the insular to both attentional processing and the symptoms of PSD. Damage to this region may not only impair cognitive function but may also predispose patients to depression, potentially through disrupted integration of cognitive and emotional processes ^38^. This supports the view that the insula serves as a key hub where cognitive dysfunction and mood disturbance converge in the aftermath of stroke.

Previous studies have consistently shown that PSD is accompanied by cognitive impairments, with emphasis on deficits in attention and executive functions in relation to depressive symptom after stroke ^6,8-10^. The PCA findings are consistent with these studies, highlighting the significant concurrence between attentional deficits and depressive symptoms in PSD. Importantly, our results extend prior work by providing further neuroanatomical evidence that directly links attentional deficits to structural damage in the insular cortex in patients with PSD.

The insular cortex has been recognized as a core hub integrating interoceptive awareness, emotional processing, and cognitive control ^20^. Structural and functional alterations in the insula have been frequently implicated in MDD ^23,25^. In particular, the left dorsal anterior insula has been shown to support attention and executive control in healthy and clinical populations ^39^. This region is also a key node in the salience network, mediating interactions between the fronto‐parietal attention network and ventral attention systems during task engagement ^40^. In MDD, reduced grey matter volume and impaired functional connectivity in the anterior insula are commonly observed and associated with deficits in attentional and emotional regulation ^18,22^. Our findings extend this association to PSD, highlighting the insula’s central role in mediating both attentional deficits and depressive symptoms following stroke. The observation of more extensive insular lesions in the PSD group suggests a relationship between the severity of insular damage and depressive symptoms, potentially contributing to sustained cognitive impairment.

While most prior work has emphasized the anterior insula, the present results point to a possible role of the posterior insula. Unlike the anterior insula, which is strongly linked to higher-order cognitive and emotional control, the posterior insula is primarily involved in sensory integration, somatosensory awareness, and visceral-autonomic processing ^41,42^. It serves as an interface between bodily states and higher-level emotional and cognitive processes, relaying interoceptive and nociceptive signals to anterior regions. Lesions in this region may therefore disrupt the integration of sensory and interoceptive information with emotional evaluation, weakening the capacity to regulate affective responses. Moreover, the posterior insula has been implicated in attention reorientation and network switching, processes that are critical for maintaining goal-directed behavior in the presence of internal or external distractions ^43^. Group difference in the posterior insula suggest that damage in this region may impair the coordination between interoceptive signals and attentional control mechanisms, thereby exacerbating both cognitive deficits and depressive symptoms in PSD. This aligns with the view that the insula, spanning anterior to posterior subregions, functions as a gradient hub with the posterior insula anchoring basic sensory-interoceptive functions that feed into the anterior insula’s role in salience detection, attention, and emotional regulation ^44^.

This study has several limitations, most notably the relatively small sample size. Stroke is characterized by substantial heterogeneity in lesion location, type, and clinical presentation, which makes it challenging to predict functional outcomes across physical, cognitive, and emotional domains ^45,46^. To generalize our findings, future research should aim to include larger and more homogeneous populations, stratified by factors such as lesion site (cortical vs. subcortical), stroke type (ischemic vs. hemorrhagic), and stroke stage (acute vs. chronic). PSD is most commonly observed within the first three months after onset, and its occurrence is strongly linked to functional outcomes and rehabilitation trajectories ^2^. The neuroanatomical correlates of PSD may differ between the acute stage and chronic PSD, reflecting distinct mechanisms across the course of recovery. Despite these limitations, the present study makes a valuable contribution as one of the first attempts to examine the relationship between cognitive function and its neural correlates in PSD, offering insights that may guide future, larger-scale investigations.

Understanding the neural underpinnings of PSD, particularly in relation to cognitive impairments, has critical implications for the development of individualized rehabilitation strategies. It remains unclear whether depression hampers rehabilitation due to severe mood disturbances and loss of motivation, or whether slower functional recovery itself contributes to depressive symptoms. Future research is therefore to define the cause and the subtypes of PSD. Longitudinal studies combined with functional imaging will be valuable in elucidating the dynamic interplay between lesion characteristics, cognitive recovery, and emotional regulation.

In conclusion, PSD presents a significant mood disorder following stroke, accompanying with attentional deficits compared to non-depressive stroke patients. The present findings suggest that the insula plays a crucial role in mediating attentional networks that are closely linked to the depressive symptoms in patients with stroke.

## Data availability

The data used in this study are available upon reasonable request from the corresponding authors. The data are not publicly available as they include information that may compromise participant confidentiality.

## Patient consent

This study was conducted with the approval by the Institutional Review Board (IRB) committee at Korean University Anam Hospital (IRB no. 2025AN0050). As this study was conducted restrospectively, the requirement for informed consent from patients was waived.

## Reference

1. Gaete JM, Bogousslavsky J. Post-stroke depression. Expert Review of Neurotherapeutics. 2008;8(1):75–92. doi:10.1586/14737175.8.1.75

2. Butsing N, Zauszniewski JA, Ruksakulpiwat S, Griffin MTQ, Niyomyart A. Association between post-stroke depression and functional outcomes: A systematic review. PLOS ONE. 2024;19(8):e0309158. doi:10.1371/journal.pone.0309158

3. Shi Y, Yang D, Zeng Y, Wu W. Risk Factors for Post-stroke Depression: A Meta-analysis. Frontiers in Aging Neuroscience. 2017;9 doi:10.3389/fnagi.2017.00218

4. Shewangizaw S, Fekadu W, Gebregzihabhier Y, Mihretu A, Sackley C, Alem A. Impact of depression on stroke outcomes among stroke survivors: Systematic review and meta-analysis. PLOS ONE. 2023;18(12):e0294668. doi:10.1371/journal.pone.0294668

5. Liu L, Marshall IJ, Li X, et al. Long-term outcomes of depression up to 10-years after stroke in the South London Stroke Register: a population-based study. The Lancet Regional Health - Europe. 2025;54:101324. doi:10.1016/j.lanepe.2025.101324

6. Kauhanen ML, Korpelainen JT, Hiltunen P, et al. Poststroke Depression Correlates With Cognitive Impairment and Neurological Deficits. Stroke. 1999;30(9):1875–1880. doi:10.1161/01.str.30.9.1875

7. Robinson RG, Jorge RE. Post-stroke depression: a review. American Journal of Psychiatry. 2016;173(3):221–231.

8. Rock PL, Roiser JP, Riedel WJ, Blackwell AD. Cognitive impairment in depression: a systematic review and meta-analysis. Psychological Medicine. 2014;44(10):2029–2040. doi:10.1017/s0033291713002535

9. McDermott LM, Ebmeier KP. A meta-analysis of depression severity and cognitive function. Journal of Affective Disorders. 2009/12/01/ 2009;119(1):1–8. doi:10.1016/j.jad.2009.04.022

10. Scult MA, Paulli AR, Mazure ES, Moffitt TE, Hariri AR, Strauman TJ. The association between cognitive function and subsequent depression: a systematic review and meta-analysis. Psychological Medicine. 2017;47(1):1–17. doi:10.1017/s0033291716002075

11. Terroni L, Sobreiro MFM, Conforto AB, et al. Association among depression, cognitive impairment and executive dysfunction after stroke. Dementia & Neuropsychologia. 2012;6(3):152–157. doi:10.1590/s1980-57642012dn06030007

12. Kang C. Predictors of Post-stroke Cognition Among Geriatric Patients: The Role of Demographics, Pre-stroke Cognition, and Trajectories of Depression. Frontiers in Psychology. 2021;12 doi:10.3389/fpsyg.2021.717817

13. Robinson RG, Starr LB, Kubos KL, Price TR. A two-year longitudinal study of post-stroke mood disorders: findings during the initial evaluation. Stroke. 1983;14(5):736–741. doi:10.1161/01.str.14.5.736

14. Uwajeh K, Egbuchulem KI, Afolabi OA. MAJOR DEPRESSIVE DISORDER: COGNITIVE, EMOTIVE AND MOTIVATIONAL CONSEQUENCES IN ADOLESCENTS. Ann Ib Postgrad Med. Apr 30 2024;22(1):116–120.

15. Zheng K, Liu Z, Miao Z, et al. Impaired cognitive flexibility in major depressive disorder: Evidences from spatial-temporal ERPs analysis. Journal of Affective Disorders. 2024/11/15/ 2024;365:406–416. doi:10.1016/j.jad.2024.08.092

16. Zhou J, Xu J, Feng Z, et al. A longitudinal analysis of the relationship between emotional symptoms and cognitive function in patients with major depressive disorder. Psychological Medicine. 2025;55 doi:10.1017/s0033291725001011

17. Sprengelmeyer R, Steele JD, Mwangi B, et al. The insular cortex and the neuroanatomy of major depression. Journal of affective disorders. 2011;133(1-2):120–127.

18. Yan F, Zan S, Xu J, Zhao S, Wang Z, Yang F. Cognitive and clinical dimensions of structural and functional insula alterations in patients with depression: a resting-state fMRI study. International Journal of Neuroscience. 2025:1–10. doi:10.1080/00207454.2024.2446908

19. Yulug B, Yalcinkaya A, Safa SS, et al. Subjective cognitive decline in major depressive patients is associated with altered entropy and connectivity changes of temporal and insular region. Translational Psychiatry. 2025;15(1) doi:10.1038/s41398-025-03518-w

20. Seeley WW, Menon V, Schatzberg AF, et al. Dissociable Intrinsic Connectivity Networks for Salience Processing and Executive Control. The Journal of Neuroscience. 2007;27(9):2349–2356. doi:10.1523/jneurosci.5587-06.2007

21. Stratmann M, Konrad C, Kugel H, et al. Insular and Hippocampal Gray Matter Volume Reductions in Patients with Major Depressive Disorder. PLoS ONE. 2014;9(7):e102692. doi:10.1371/journal.pone.0102692

22. Sliz D, Hayley S. Major Depressive Disorder and Alterations in Insular Cortical Activity: A Review of Current Functional Magnetic Imaging Research. Frontiers in Human Neuroscience. 2012;6 doi:10.3389/fnhum.2012.00323

23. Schnellbächer GJ, Rajkumar R, Veselinović T, et al. Structural alterations of the insula in depression patients – A 7-Tesla-MRI study. NeuroImage: Clinical. 2022/01/01/ 2022;36:103249. doi:10.1016/j.nicl.2022.103249

24. Lynch CJ, Elbau IG, Ng T, et al. Frontostriatal salience network expansion in individuals in depression. Nature. 2024;633(8030):624–633. doi:10.1038/s41586-024-07805-2

25. Avery JA, Drevets WC, Moseman SE, Bodurka J, Barcalow JC, Simmons WK. Major Depressive Disorder Is Associated With Abnormal Interoceptive Activity and Functional Connectivity in the Insula. Biological Psychiatry. 2014;76(3):258–266. doi:10.1016/j.biopsych.2013.11.027

26. Shi Y, Zeng Y, Wu L, et al. A Study of the Brain Abnormalities of Post-Stroke Depression in Frontal Lobe Lesion. Scientific Reports. 2017;7(1) doi:10.1038/s41598-017-13681-w

27. Klingbeil J, Brandt M-L, Stockert A, et al. Associations of lesion location, structural disconnection, and functional diaschisis with depressive symptoms post stroke. Frontiers in Neurology. 2023;14 doi:10.3389/fneur.2023.1144228

28. Beck AT, Ward C, Mendelson M, Mock J, Erbaugh J. Beck depression inventory (BDI). Arch gen psychiatry. 1961;4(6):561–571.

29. Lasa L, Ayuso-Mateos JL, Vázquez-Barquero JL, Díez-Manrique FJ, Dowrick CF. The use of the Beck Depression Inventory to screen for depression in the general population: a preliminary analysis. J Affect Disord. Jan-Mar 2000;57(1-3):261–5. doi:10.1016/s0165-0327(99)00088-9

30. Leentjens AFG, Verhey FRJ, Luijckx G-J, Troost J. The validity of the Beck Depression Inventory as a screening and diagnostic instrument for depression in patients with Parkinson’s disease. Movement Disorders. 2000;15(6):1221–1224. doi:10.1002/1531-8257(200011)15:6<1221::aid-mds1024>3.0.co;2-h

31. Hedayati SS, Bosworth HB, Kuchibhatla M, Kimmel PL, Szczech LA. The predictive value of self-report scales compared with physician diagnosis of depression in hemodialysis patients. Kidney International. 2006;69(9):1662–1668. doi:10.1038/sj.ki.5000308

32. Bai D, Lee JB, Bahn YK. Computerized Neurocognitive Function Test. Seoul: Hana Medical Publishing Co. 2005:12–20.

33. Ha K-S, Kwon JS, Lyoo I-K, Kong SW, Lee DW, Youn T. Development and Standardization Process, and Factor Analysis of the Computerized Cognitive Function Test System for Korea Adults. J Korean Neuropsychiatr Assoc. 2002;41(3)

34. Lyoo I-K, Kwon JS, Ha K-S. Development and Standardization of the Computerized Higher Cortical Function Assessment for Korean Adults. J Korean Neuropsychiatr Assoc. 2002;41(3)

35. Abdi H, Williams LJ. Principal component analysis. WIREs Computational Statistics. 2010;2(4):433–459. doi:10.1002/wics.101

36. Yushkevich PA, Piven J, Hazlett HC, et al. User-guided 3D active contour segmentation of anatomical structures: Significantly improved efficiency and reliability. NeuroImage. 2006;31(3):1116–1128. doi:10.1016/j.neuroimage.2006.01.015

37. Amunts K, Mohlberg H, Bludau S, Zilles K. Julich-Brain: A 3D probabilistic atlas of the human brain’s cytoarchitecture. Science. 2020;369(6506):988–992. doi:10.1126/science.abb4588

38. Namkung H, Kim S-H, Sawa A. The Insula: An Underestimated Brain Area in Clinical Neuroscience, Psychiatry, and Neurology. Trends in Neurosciences. 2017;40(4):200–207. doi:10.1016/j.tins.2017.02.002

39. Nelson SM, Dosenbach NUF, Cohen AL, Wheeler ME, Schlaggar BL, Petersen SE. Role of the anterior insula in task-level control and focal attention. Brain Structure and Function. 2010;214(5-6):669–680. doi:10.1007/s00429-010-0260-2

40. Menon V, Uddin LQ. Saliency, switching, attention and control: a network model of insula function. Brain structure and function. 2010;214(5):655–667.

41. Uddin LQ, Nomi JS, Hébert-Seropian B, Ghaziri J, Boucher O. Structure and Function of the Human Insula. Journal of Clinical Neurophysiology. 2017;34(4):300–306. doi:10.1097/wnp.0000000000000377

42. Craig AD. How do you feel — now? The anterior insula and human awareness. Nature Reviews Neuroscience. 2009;10(1):59–70. doi:10.1038/nrn2555

43. Cauda F, Costa T, Torta DME, et al. Meta-analytic clustering of the insular cortex. NeuroImage. 2012;62(1):343–355. doi:10.1016/j.neuroimage.2012.04.012

44. Centanni SW, Janes AC, Haggerty DL, Atwood B, Hopf FW. Better living through understanding the insula: Why subregions can make all the difference. Neuropharmacology. 2021/10/15/ 2021;198:108765. doi:10.1016/j.neuropharm.2021.108765

45. Kent TA, Soukup VM, Fabian RH. Heterogeneity Affecting Outcome From Acute Stroke Therapy. Stroke. 2001;32(10):2318–2327. doi:10.1161/hs1001.096588

46. Gao MM, Wang J, Saposnik G. The Art and Science of Stroke Outcome Prognostication. Stroke. 2020;51(5):1358–1360. doi:10.1161/strokeaha.120.028980

